# Protocol: A two-wave cross-sectional study in England investigating suicidal behaviour and self-harm amongst healthcare workers during the Covid-19 pandemic

**DOI:** 10.1101/2021.09.16.21263255

**Authors:** Prianka Padmanathan, Danielle Lamb, Hannah Scott, Simon Wessely, Paul Moran

## Abstract

**Introduction:** There have been longstanding concerns regarding an increased risk of suicide amongst healthcare workers. The Covid-19 pandemic has placed an additional burden on staff, yet few studies have investigated the impact of the pandemic on their risk of suicide and self-harm. We aimed to investigate the cumulative incidence, prevalence, and correlates of suicidal ideation, suicide attempts, and non-suicidal self-injury amongst healthcare workers during the Covid-19 pandemic.

**Methods and Analysis:** NHS Check is an online survey that was distributed to all staff (clinical and non-clinical), students, and volunteers in 18 NHS Trusts across England during the Covid-19 pandemic. Data collected in wave 1 (collected between April 2020 and January 2021) and wave 2 (collected 6 months after wave 1) will be analysed. The full cohort of wave 1 participants will be weighted to represent the age, sex, ethnicity, and roles profile of the workforce at each Trust, and the weighted prevalence and cumulative incidence of suicidal ideation, suicide attempts, and non-suicidal self-injury will be described. Two-level random effects logistic regression models will be used to investigate the relationship between suicidal behaviour and self-harm, and demographic characteristics (age, sex, ethnicity) and workplace factors (concerns regarding access to personal protective equipment, re-deployment status, moral injury, confidence around raising and the management of safety concerns, support by supervisors or managers, satisfaction with standard of care provided). Results will be stratified by role (clinical/non-clinical).

**Changes in this protocol compared with the original version:**

1. All variables describing workplace factors will be analysed as binary variables for consistency.
2. The responses to two questions on raising, and the management of, safety concerns will be analysed separately rather than combined to minimise loss of information.
3. Binary suicide-related outcomes will be used in the logistic regression analyses, where the presence of an outcome represents its occurrence within the previous one (wave 2) or two months (wave 1), specifically. This was previously not specified.

## Introduction

There have been longstanding concerns regarding an increased risk of suicide amongst healthcare workers ^1,2^. The Covid-19 pandemic has placed an additional burden on healthcare staff, yet while several studies have investigated the impact of the pandemic on their mental health ^3-7^, few have investigated their risk of suicide and self-harm ^8^. Provisional Office of National Statistics data indicate that there has been an increase in the absolute number of suicides, and the proportion of total deaths that were due to suicide, amongst health workers in England and Wales in the first six months of 2020 compared to the same period in 2019 ^9^. Occupational exposures such as inadequate support from colleagues and moral injury have previously been linked with suicidal ideation and suicide attempts^10^, although, to date, the latter has only been investigated in a small number of studies of military personnel ^11^. Amongst healthcare workers working in epidemics, several modifiable occupational risk factors, such as a lack of specialised training or personal protective equipment, have been associated with worse mental health ^6,12^.

## Aims & objectives

To investigate the cumulative incidence, prevalence, and correlates of non-fatal suicidal behaviour (i.e. suicidal ideation and suicide attempts) and self-harm (i.e. non-suicidal self-injury) amongst healthcare workers during the Covid-19 pandemic.

1. Describe the prevalence of suicidal ideation, suicide attempts, and non-suicidal self-injury at baseline and 6 months follow-up
2. Describe the cumulative incidence of suicidal ideation, suicide attempts, and non-suicidal self-injury at 6 months follow-up
3. Investigate the relationship between demographic characteristics and workplace factors reported at baseline and suicidal ideation, suicide attempts, and self-harm at baseline and at 6 months follow-up.

## Methods

NHS Check is a multi-wave online survey that was distributed to all staff (clinical and non-clinical), students, and volunteers in 18 NHS Trusts across England during the Covid-19 pandemic ^13^. Data from the 17 Trusts (those with a response rate of over 5%) will be included in this analysis. Data for wave 1 were collected between April 2020 and January 2021. Data for wave 2 were collected six months after each participant’s completion of the wave 1 survey. Both waves included a “short” survey and a “long” survey. Participants could choose to complete either the “short” survey alone or complete the “long” survey in addition. In wave 1, the “short” survey collected demographic information, whilst the “long” survey included questions relating to the workforce factors and the suicide and self-harm outcomes described in this study. In wave 2, questions relating to suicide and self-harm were included in the “short” survey. Further details regarding data collection can be found elsewhere ^13^.

### Outcomes

Suicidal ideation, suicide attempts and non-suicidal self-injury were assessed using the following items from the Clinical Interview Schedule (CIS-R)^14^: “Have you ever thought of taking your life, even though you would not actually do it?”; “Have you ever made an attempt to take your life, by taking an overdose of tablets or in some other way?”; and “Have you ever deliberately harmed yourself in any way but not with the intention of killing yourself?”. In the baseline survey, participants could select from the following options: “Yes, in the past 2 months”, “Yes, but not in the past 2 months”, or “No”. In the 6-month follow-up survey, participants could select from the same options but a one-month, rather than two-month, time frame was specified.

In the logistic regression analyses described below, binary suicide-related outcomes will be used, where the presence of an outcome represents its occurrence within the previous one (wave 2) or two months (wave 1), specifically.

### Explanatory factors

#### Demographic characteristics

Variables will be defined as follows based on previous NHS Check analyses ^8^:

- Age
- Sex (Female, male – options are limited by survey design)
- Ethnicity (ONS top-level categories: White, Black, Asian, Mixed ethnicity, Other ethnicity)
- Role (Doctor, nurse, other clinical, non-clinical)

#### Individual-level workplace factors

All factors will be analysed as binary exposures (see Appendix for questions and responses):

- Access to personal protective equipment (Q85 short survey: exposure=never/sometimes)
- Re-deployment status (Q69 short survey: exposure=yes)
- Moral injury (Q50 long survey: exposure=moderately agree/strongly agree to any of the sub-questions)
- Raising safety concerns (Q19 long survey: exposure=strongly disagree/disagree)
- Management of safety concerns (Q19 long survey: exposure=strongly disagree/disagree)
- Support by supervisors or managers (Q87b short survey: exposure=not at all/a little bit)
- Satisfaction with standard of care provided - worse care than usual (Q11b long survey: exposure=often/always) – clinical staff only

### Statistical analysis

The full cohort (i.e. those completing the short survey at wave 1) will be weighted using a ranking algorithm based on the age, sex, ethnicity, and roles profile of the workforce at each Trust to maximise representativeness. To complete the weighting, missing demographic data will be imputed using multiple imputation. The imputed data will only be used for the purposes of completing weighting and will not be used in any other analyses related to the aims and objectives, which will only include participants with complete data on the included variables. Some participants completed the baseline survey more than once. The weighting includes an average of these participants’ responses.

We will describe the demographic and occupational characteristics of the full cohort, our wave 1 sub-sample (i.e. those who completed the suicide and self-harm questions in the wave 1 long survey), and our wave 2 sub-sample (i.e. those who completed the suicide and self-harm questions in the wave 2 short survey) (Table 1). We will compare our sub-samples to the full cohort, using Chi^2^ tests for each binary or categorical variable, to assess any statistically significant differences between the groups.

**Table 1:** Survey sample characteristics

| Variable | Category | No. (%) unless otherwise specified |  |  |
| --- | --- | --- | --- | --- |
|  |  | Full cohort<br>(n=) | Wave 1 cohort<br>(n=) | Wave 2 cohort<br>(n=) |
| Age in years<sup>α</sup> | ≤30 |  |  |  |
|  | 31-40 |  |  |  |
|  | 41-50 |  |  |  |
|  | 51-60 |  |  |  |
|  | ≥61 |  |  |  |
|  | Missing |  |  |  |
| Sex<sup>α</sup> | Female |  |  |  |
|  | Male |  |  |  |
|  | Missing |  |  |  |
| Ethnicity<sup>α</sup> | White |  |  |  |
|  | Black |  |  |  |
|  | Asian |  |  |  |
|  | Mixed/Multiple racial & ethnic groups |  |  |  |
|  | Other racial & ethnic minority groups |  |  |  |
|  | Missing |  |  |  |
| Role<sup>α</sup> | Doctor |  |  |  |
|  | Nurse |  |  |  |
|  | Other clinical |  |  |  |
|  | Non-clinical |  |  |  |
|  | Missing |  |  |  |
| Redeployment<sup>α</sup> | No |  |  |  |
|  | Yes |  |  |  |
|  | Missing |  |  |  |
| Concerns about access to PPE<sup>αγ</sup> | No |  |  |  |
|  | Yes |  |  |  |
|  | Missing |  |  |  |
| Concerns about support from managers<sup>α</sup> | *No* |  |  |  |
|  | *Yes* |  |  |  |
|  | *Missing* |  |  |  |
| Concerns about raising safety concerns<sup>βγ</sup> | *No* |  |  |  |
|  | *Yes* |  |  |  |
|  | *Missing* |  |  |  |
| Concerns about the management of safety concerns<sup>βγ</sup> | *No* |  |  |  |
|  | *Yes* |  |  |  |
|  | *Missing* |  |  |  |
| Dissatisfaction with standard of care provided<sup>βγ</sup> | *No* |  |  |  |
|  | *Yes* |  |  |  |
|  | *Missing* |  |  |  |
| Moral injury<sup>βγ</sup> | *No* |  |  |  |
|  | *Yes* |  |  |  |
|  | *Missing* |  |  |  |

We will describe the weighted prevalence of our outcomes for participants at wave 1 and at wave 2 (Table 2). Due to the discrepancy in the time frames specified by the suicide and self-harm questions between waves, a 2-month period prevalence will be calculated for wave 1, whilst a 1-month period prevalence will be calculated for wave 2. We will also describe the weighted cumulative incidence of our outcomes at wave 2 i.e. the number and percentage of participants who responded “no” to previous self-harm and suicidal behaviour at wave 1, but “yes” at wave 2 (Table 2).

**Table 2:** Prevalence and incidence of suicidal-related outcomes

|  |  |  | Prevalence* |  |  | Cumulative incidence |  |  |
| --- | --- | --- | --- | --- | --- | --- | --- | --- |
|  | Response |  | Suicidal ideation | Suicidal attempts | Non-suicidal self-injury | Suicidal ideation | Suicidal attempts | Non-suicidal self-injury |
| Wave 1 | No | n (%)<br>[95%CI] |  |  |  | N/a | N/a | N/a |
|  | Yes, but not in the 2 months prior to wave 1 survey completion | n (%)<br>[95%CI] |  |  |  | N/a | N/a | N/a |
|  | Yes, in the 2 months prior to wave 1 survey completion | n (%)<br>[95%CI] |  |  |  | N/a | N/a | N/a |
| Wave 2 | No | n (%)<br>[95%CI] |  |  |  |  |  |  |
|  | Yes, but not in the month prior to wave 2 survey completion | n (%)<br>[95%CI] |  |  |  |  |  |  |
|  | Yes, in the month prior to wave 2 survey completion | n (%)<br>[95%CI] |  |  |  |  |  |  |

n=frequency (unweighted raw data); percentages are weighted
\*Wave 1=2-month period prevalence; Wave 2=1-month period prevalence

Given the clustered nature of the data, we will then use two-level random effects logistic regression models, to investigate the relationship between each demographic characteristic and workplace factor reported at wave 1 and suicidal behaviour and self-harm at wave 1 and wave 2 (Tables 3 and 4). Data will be stratified by role (clinical/non-clinical). Participants (level 1) will be clustered within each Trust (level 2), and we will adjust for age, sex, ethnicity and date of survey completion as level 1 covariates. When analysing the outcomes at wave 2, in addition, we will also adjust for the corresponding outcome at wave 1 (level 1). For example, when analysing suicidal ideation at wave 2, we will adjust for suicidal ideation at wave 1. In each model, we will only include participants with complete data on all included variables. We will use the “subpop” command in Stata to ensure that the standard errors of the estimates are calculated correctly ^16^.

**Table 3:** Association between demographic characteristics and suicide-related outcomes at wave 1 and wave 2

| Variable | Category | Wave 1 |  |  |  |  |  | Wave 2 |  |  |  |  |  |
| --- | --- | --- | --- | --- | --- | --- | --- | --- | --- | --- | --- | --- | --- |
|  |  | Suicidal ideation<br>(aOR; 95% CI) |  | Suicide<br>attempts<br>(aOR; 95% CI) |  | Non-suicidal<br>self-injury<br>(aOR; 95% CI) |  | Suicidal ideation<br>(aOR; 95% CI) |  | Suicide<br>attempts<br>(aOR; 95% CI) |  | Non-suicidal<br>self-injury<br>(aOR; 95% CI) |  |
|  |  | Clinical | Non-clinical | Clinical | Non-clinical | Clinical | Non-clinical | Clinical | Non-clinical | Clinical | Non-clinical | Clinical | Non-clinical |
| Age in<br>years |  |  |  |  |  |  |  |  |  |  |  |  |  |
| Sex | Female (Ref) |  |  |  |  |  |  |  |  |  |  |  |  |
|  | Male |  |  |  |  |  |  |  |  |  |  |  |  |
| Ethnicity | White (Ref) |  |  |  |  |  |  |  |  |  |  |  |  |
|  | Black/African/Caribbean/Black<br>British |  |  |  |  |  |  |  |  |  |  |  |  |
|  | Asian/Asian British |  |  |  |  |  |  |  |  |  |  |  |  |
|  | Mixed/Multiple racial and<br>ethnic groups |  |  |  |  |  |  |  |  |  |  |  |  |
|  | Other racial and ethnic<br>minority groups |  |  |  |  |  |  |  |  |  |  |  |  |

aOR: adjusted odds ratios: adjusted for age, sex, ethnicity, and date of survey completion (6-month data also adjusted for corresponding outcome measured at baseline); CI: confidence intervals

**Table 4:** Association between baseline workplace factors and suicide-related outcomes at wave 1 and wave 2

| Variable | Category | Wave 1 survey |  |  |  |  |  | Wave 2 survey |  |  |  |  |  |
| --- | --- | --- | --- | --- | --- | --- | --- | --- | --- | --- | --- | --- | --- |
|  |  | Suicidal ideation<br>(aOR; 95% CI) |  | Suicide attempts<br>(aOR; 95% CI) |  | Non-suicidal self-injury<br>(aOR; 95% CI) |  | Suicidal ideation<br>(aOR; 95% CI) |  | Suicide attempts<br>(aOR; 95% CI) |  | Non-suicidal self-injury<br>(aOR; 95% CI) |  |
|  |  | Clinical | Non-clinical | Clinical | Non-clinical | Clinical | Non-clinical | Clinical | Non-clinical | Clinical | Non-clinical | Clinical | Non-clinical |
| Redeployment | No (Ref) |  |  |  |  |  |  |  |  |  |  |  |  |
|  | Yes |  |  |  |  |  |  |  |  |  |  |  |  |
| Concerns around raising safety issues | No (Ref) |  |  |  |  |  |  |  |  |  |  |  |  |
|  | Yes |  |  |  |  |  |  |  |  |  |  |  |  |
| Concerns around management of safety issues | No (Ref) |  |  |  |  |  |  |  |  |  |  |  |  |
|  | Yes |  |  |  |  |  |  |  |  |  |  |  |  |
| Concerns about access to PPE | No (Ref) |  |  |  |  |  |  |  |  |  |  |  |  |
|  | Yes |  |  |  |  |  |  |  |  |  |  |  |  |
| Concerns about support from managers | No (Ref) |  |  |  |  |  |  |  |  |  |  |  |  |
|  | Yes |  |  |  |  |  |  |  |  |  |  |  |  |
| Concerns about standard of care provided | No (Ref) |  |  |  |  |  |  |  |  |  |  |  |  |
|  | Yes |  |  |  |  |  |  |  |  |  |  |  |  |
| Moral injury | No (Ref) |  |  |  |  |  |  |  |  |  |  |  |  |
|  | Yes |  |  |  |  |  |  |  |  |  |  |  |  |

\*aOR: adjusted odds ratios – adjusted for age, sex & ethnicity, and date of survey completion (6-month data also adjusted for corresponding outcome measured at wave 1); CI: confidence intervals

### Ethics

Ethics and Dissemination Ethical approval for the NHS Check study was granted by the Health Research Authority (reference: 20/HRA/210, IRAS: 282686) and local Trust Research and Development approval. See elsewhere for further details ^13^.

## Data Availability

Access to study data by researchers may be possible on application to the chief investigators of NHS Check.

## Acknowledgements

We wish to acknowledge the National Institute of Health Research (NIHR) Applied Research Collaboration (ARC) National NHS and Social Care Workforce Group, with the following ARCs: East Midlands, East of England, South West Peninsula, South London, West, North West Coast, Yorkshire and Humber, and North East and North Cumbria. They enabled the set-up of the national network of participating hospital sites and aided the research team to recruit effectively during the COVID-19 pandemic.

The NHS CHECK consortium includes the following site leads: Sean Cross, Amy Dewar, Chris Dickens, Mary Docherty, Frances Farnworth, Adam Gordon, Charles Goss, Jessica Harvey, Nusrat Husain, Peter Jones, Damien Longson, Richard Morriss, Jesus Perez, Mark Pietroni, Ian Smith, Tayyeb Tahir, Peter Trigwell, Jeremy Turner, Julian Walker, Scott Weich, Ashley Wilkie.

The NHS CHECK consortium includes the following co-investigators and collaborators: Peter Aitken, Anthony David, Sarah Dorrington, Rosie Duncan, Cerisse Gunasinghe, Stephani Hatch, Daniel Leightley, Ira Madan, Isabel McMullen, Martin Parsons, Paul Moran, Dominic Murphy, Catherine Polling, Alexandra Pollitt, Danai Serfioti, Chloe Simela, Charlotte Wilson Jones.

