## Appendix for "Protocol: A two-wave cross-sectional study in England investigating suicidal behaviour and self-harm amongst healthcare workers during the Covid-19 pandemic"

Survey questions relating to individual level workplace factors (answers in red to be coded as the exposure):

Q11b (long survey): Satisfaction with standard of care provided

In the past 2 weeks I or my team have had to provide significantly worse care than usual to our patients or deny them treatment that would normally be available

(Answer options: Never, Rarely, Sometimes, Often, Always, Not applicable)

Q19 (long survey): Confidence around raising and the management of safety concerns

Raising concerns about unsafe clinical practice:

a) I would feel secure raising concerns about unsafe clinical practice.
b) I am confident that my organisation would address my concern.

(Answer options: Strongly disagree, Disagree, Neither agree nor disagree, Agree, Strongly agree, Not applicable)

Q50 (long survey): Moral injury event scale

Please select how much you agree or disagree with each of the following statements regarding your experiences at any time since working for/with the NHS during COVID-19 (coronavirus) pandemic:

a) I saw things that were morally wrong
b) I am troubled by having witnessed others’ immoral acts
c) I acted in ways that violated my own moral code or values
d) I am troubled by having acted in ways that violated my own morals or values
e) I violated my own morals by failing to do something that I felt I should have done
f) I am troubled because I violated my morals by failing to do something that I felt I should have done
g) I feel betrayed by my supervisors/managers who I once trusted
h) I feel betrayed by co-workers who I once trusted
i) I feel betrayed by others outside the health service who I once trusted

(Answer options: Strongly disagree, Moderately disagree, Slightly disagree, Slightly agree, Moderately agree, Strongly agree, Not applicable)

Q69 (short survey): Re-deployment status

Prior to the COVID-19 (coronavirus) pandemic did you work in a different setting?

(Answer options: Yes, No)

Q85 (short survey): Access to personal protective equipment

Do you have access to adequate personal protective equipment (PPE)?

(Answer options: Never, Sometimes, Often, Always, Not applicable)

Q87b (short survey): Support by supervisors or managers

Since the COVID-19 (coronavirus) pandemic how well do you feel supported by your supervisors/managers?

(Answer options: Not at all, A little bit, Moderately, Quite a bit, Extremely)
